# Precision Phenotyping for Curating Research Cohorts of Patients with Post-Acute Sequelae of COVID-19 (PASC) as a Diagnosis of Exclusion

**DOI:** 10.1101/2024.04.13.24305771

**Authors:** Alaleh Azhir, Jonas Hügel, Jiazi Tian, Jingya Cheng, Ingrid V. Bassett, Douglas S. Bell, Elmer V. Bernstam, Maha R. Farhat, Darren W. Henderson, Emily S. Lau, Michele Morris, Yevgeniy R. Semenov, Virginia A. Triant, Shyam Visweswaran, Zachary H. Strasser, Jeffrey G. Klann, Shawn N. Murphy, Hossein Estiri

## Abstract

Scalable identification of patients with the post-acute sequelae of COVID-19 (PASC) is challenging due to a lack of reproducible precision phenotyping algorithms and the suboptimal accuracy, demographic biases, and underestimation of the PASC diagnosis code (ICD-10 U09.9). In a retrospective case-control study, we developed a precision phenotyping algorithm for identifying research cohorts of PASC patients, defined as a diagnosis of exclusion. We used longitudinal electronic health records (EHR) data from over 295 thousand patients from 14 hospitals and 20 community health centers in Massachusetts. The algorithm employs an attention mechanism to exclude sequelae that prior conditions can explain. We performed independent chart reviews to tune and validate our precision phenotyping algorithm. Our PASC phenotyping algorithm improves precision and prevalence estimation and reduces bias in identifying Long COVID patients compared to the U09.9 diagnosis code. Our algorithm identified a PASC research cohort of over 24 thousand patients (compared to about 6 thousand when using the U09.9 diagnosis code), with a 79.9 percent precision (compared to 77.8 percent from the U09.9 diagnosis code). Our estimated prevalence of PASC was 22.8 percent, which is close to the national estimates for the region. We also provide an in-depth analysis outlining the clinical attributes, encompassing identified lingering effects by organ, comorbidity profiles, and temporal differences in the risk of PASC. The PASC phenotyping method presented in this study boasts superior precision, accurately gauges the prevalence of PASC without underestimating it, and exhibits less bias in pinpointing Long COVID patients. The PASC cohort derived from our algorithm will serve as a springboard for delving into Long COVID’s genetic, metabolomic, and clinical intricacies, surmounting the constraints of recent PASC cohort studies, which were hampered by their limited size and available outcome data.

## Introduction

The COVID-19 pandemic exerted widespread, long-lasting impacts on the full spectrum of human health. As we move into the endemic phase of SARS-CoV-2, a considerable proportion of the population continues to struggle with prolonged symptoms following their initial exposure to SARS-CoV-2. Post-acute sequelae of SARS-CoV-2/COVID-19 (PASC), also referred to as Post-COVID Conditions (PCC) and Long COVID,^1,2^ has emerged as a complex issue, subject to ongoing scientific and political debate globally. A growing body of evidence suggests that post-acute sequelae of SARS-CoV-2 infection affect multi-organ systems.^3–9^

Despite extensive efforts to characterize and evaluate risks for PASC, a scalable, universally accepted algorithm for identifying patients who may be suffering from PASC is lacking. In the U.S., the current diagnostic codes (e.g., ICD-10 code, U09. 9: Post COVID-19 condition) lack sensitivity and specificity for accurately identifying afflicted patients.^47–50^ Zhang et al. (2023)^51^ demonstrated that the U09. 9 (ICD-10 code) can be an untrustworthy proxy for gauging long-COVID, where the positive predictive value (PPV) ranged from 40 to 65 percent, based on the PASC reference definition. In addition, Pfaff et al. (2023) found a notable tilt in the demographic makeup of patients diagnosed with U09.9 towards women, White, non-Hispanic individuals, along with those residing in regions characterized by low poverty rates, high educational attainment, and ample access to medical services.^48^ The lack of a robust phenotyping algorithm for identifying PASC patients has hindered effective enrollment for large-scale clinical studies of potential therapies.

Further, standard approaches for measuring relative effects or associations aimed at discerning conditions exhibiting elevated relative risks among an exposed group (in this case, COVID-19 patients) are insufficient to identify individuals afflicted with PASC. For instance, shortness of breath has been extensively documented as a PASC.^52^ Nevertheless, not all episodes of shortness of breath observed in individuals with a history of COVID-19 denote PASC, as such symptoms may be attributable to pre-existing conditions such as heart failure or asthma. These challenges demand particular scrutiny to enable the development of a robust algorithm for the characterization of PASC with real-world data.^51^

The World Health Organization (WHO) characterizes PASC as a diagnosis of exclusion, which offers a practical basis for identifying those suffering from PASC (henceforth referred to as long-haulers). WHO defines PASC as the continuation or development of new symptoms three months after the initial infection, lasting at least two months with no other explanation.^53,54^ Passively collected longitudinal data from electronic health records (EHRs) provide a cost-effective option for enriching cohort definitions and identifying at-risk individuals.

In this study, we introduce a reproducible precision phenotyping algorithm for the post-acute sequelae of SARS-CoV-2, based on the WHO’s definition of PASC as a diagnosis of exclusion, with clinical data from EHRs. In this case-control study, our algorithm first identifies conditions associated with SARS-CoV-2 (similar to prior studies^11,55–57^). Second, using a specialized temporal pattern mining algorithm^58^, our algorithm takes an extra step to distinguish those sequelae that cannot be explained by the patient’s past medical history. This algorithm adds a novel personalized exclusion by temporal association step to the standard risk studies, resulting in the largest validated computationally curated cohort of long-haulers. We provide a fully executable environment that can be directly applied and/or tuned to any healthcare organization with ICD-10 diagnosis and procedure codes.

Our method for identifying PASC boasts superior precision (vs. U09.9), accurately gauging the prevalence of this condition without downplaying its significance. Further, compared to the conventional U09.9 diagnosis code, our approach exhibits less bias in identifying PASC patients across demographic groups, offering a more nuanced understanding of Long COVID patients. Further, our approach enables addressing temporal questions on the recurrence and sequence of PASC following various episodes of COVID-19 infection.

In addition to introducing the precision phenotyping approach, we provide an in-depth analysis outlining the clinical attributes, encompassing identified lingering effects by organ, comorbidity profiles, and temporal differences in the risk of PASC. The comprehensive PASC cohort resulting from our precision phenotyping algorithm will enable deep dives into the multifaceted expressions of Long COVID through genetic, metabolomic, and clinical inquiries. This surpasses the constraints of earlier cohort studies, which were hampered by limited size and outcome data.

## Method

The WHO defines PASC as a diagnosis of exclusion without offering a predefined set of conditions constituting PASC – a short presentation of the methodologies can be found on GitHub: https://clai-group.github.io/long_covid_ai_implementation_guide.

We conducted a retrospective case-control study that included: (1) patients with a clinical record indicating COVID-19 infection between 03/2020 and 06/2023 (cases) and (2) a non-COVID control group from the pandemic era, and (3) a viral infection control group from the pre-pandemic era (Figure 1). Cases were patients with at least one confirmed SARS-CoV-2 infection, captured by a positive Polymerase Chain Reaction (PCR) test or a recorded diagnosis. For controls, we identified two distinct groups. The first, termed post-pandemic (post-2020) group, comprised patients with neither a record of SARS-CoV-2 infection nor any indication of probable infection. Concurrently, pre-pandemic controls included patients with a viral infection in 2018 (see Table 1S for the inclusion/exclusion criteria). We mandated at least a year of follow-up data from the last infection record for the cases and pre-pandemic controls. For cases and both controls, we extracted historical data from 3 years prior to the index date (i.e., 2017 for the pandemic cohorts and 2015 for the pre-pandemic controls).

**Table 1.** Summary statistics of the study population.

|  | COVID-19<br>Cases | Post-pandemic<br>Controls | Pre-pandemic<br>Controls |
| --- | --- | --- | --- |
| **Number of patients** | 85,364 | 170,497 | 39,817 |
| **Age Mean** | 53.6 | 53.7 | 54.7 |
| **Female Percent** | 62.6 | 62.5 | 63.2 |
| **Charlson Mean** | 2.2 | 2.2 | 2.0 |
| **Date Range** | 2017-01-01 to 2023-06-08 | 2017-01-01 to 2023-06-08 | 2015-01-01 to 2020-01-01 |
| **Race** |  |  |  |
| White | 60,935 (71.4%) | 122,646 (71.9%) | 29,687 (74.2%) |
| Other | 10,243 (12.0%) | 19,799 (11.6%) | 3,415 (8.5%) |
| Black | 8,409 (9.9%) | 16,688 (9.8%) | 3,273 (8.2%) |
| Unknown | 2,845 (3.3%) | 5,757 (3.4%) | 2,185 (5.5%) |
| Asian | 2,933 (3.4%) | 5,607 (3.3%) | 1,257 (3.1%) |
| **Hispanic ethnicity** | 6,110 (7.2%) | 8,840 (5.2%) | 1,950 (4.9%) |

**Figure 1.**
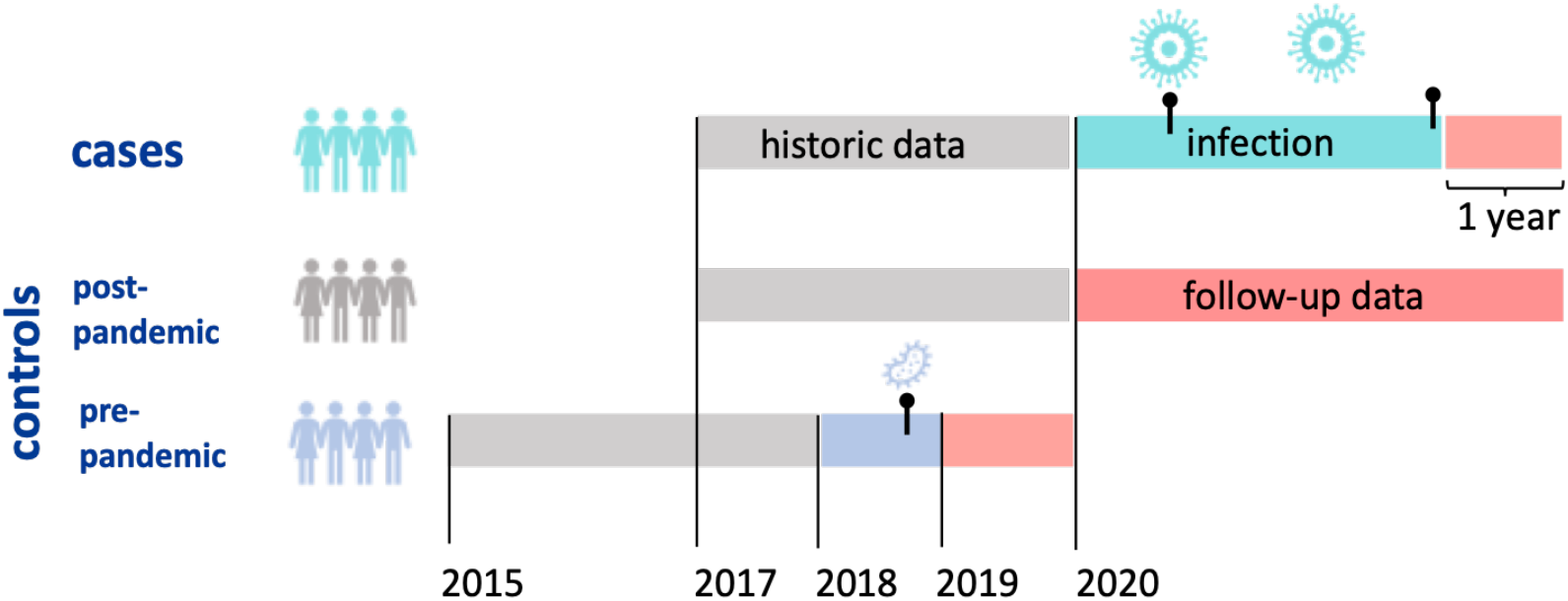
Criteria for selection of cases and controls. Cases are infected at least once by SARS-CoV-2, whereas the pre-pandemic controls had a viral infection in 2018.

### Data

We utilized electronic health record (EHR) data from 14 hospitals and 20 community health centers within the Mass General Brigham (MGB) integrated healthcare system in Massachusetts. We selected post-pandemic controls by propensity score matching them (1:2) with cases on sex, race, age, and Charlson comorbidity index scores. We extracted diagnosis and procedure data from the MGB Research Patient Data Repository^59^ and mapped them to the Clinical Classifications Software Refined (CCSR).^60^ To enrich our research cohorts by ensuring longitudinal continuity necessary for accurate inference, we utilized a validated continuity metric in which we only included a patient in our analysis if their longitudinal continuity score was above a given threshold.^61^ We also used a validated algorithm^62,63^ to temporally cluster multiple episodes of SARS-CoV-2 infections in patients with more than one infection episode. See Appendix 2 for details of matching and data harmonization.

### PASC definition

We developed a computational implementation of the WHO definition of PASC^53,54^ using a high-performance program for transitive temporal representation mining, tSPM+,^58^ previously developed for mining temporal representations from clinical data.^64^ For example, in a series of sequential events A → B → C, the transitivity property allows for mining sequence A→C (in addition to A→B and B→C). tSPM+ mines transitive sequences of medical records that begin or end with a pre-specified set of clinical concepts (i.e., CCSR categories) and are not sparse (with a given sparsity parameter). For each transitive sequence *a* → *b*, tSPM+ also computes the temporal duration, *d*_*ab*_, between the two elements of the sequence {*a, b*}, measured in months.Details of the PASC definition process are provided in Appendix 2.

To define PASC, we first identified a subset of candidate CCSR categories that meet WHO’s first two criteria – i.e., continuation or development of new symptoms after initial recovery from an acute SARS-CoV-2 episode, with these symptoms lasting for at least 2 months.

Second, the candidate PASC must be correlated with SARS-CoV-2 (obtained from the case-control design). We then developed an attention mechanism that rules out a candidate PASC if it can be explained by other prior conditions in a given patient’s medical records based on temporal associations (Spearman’s rank-order correlation^65,66^) computed from the cases and controls. Figure 2 illustrates the implementation of WHO’s PASC definition as a diagnosis of exclusion.

**Figure 2.**
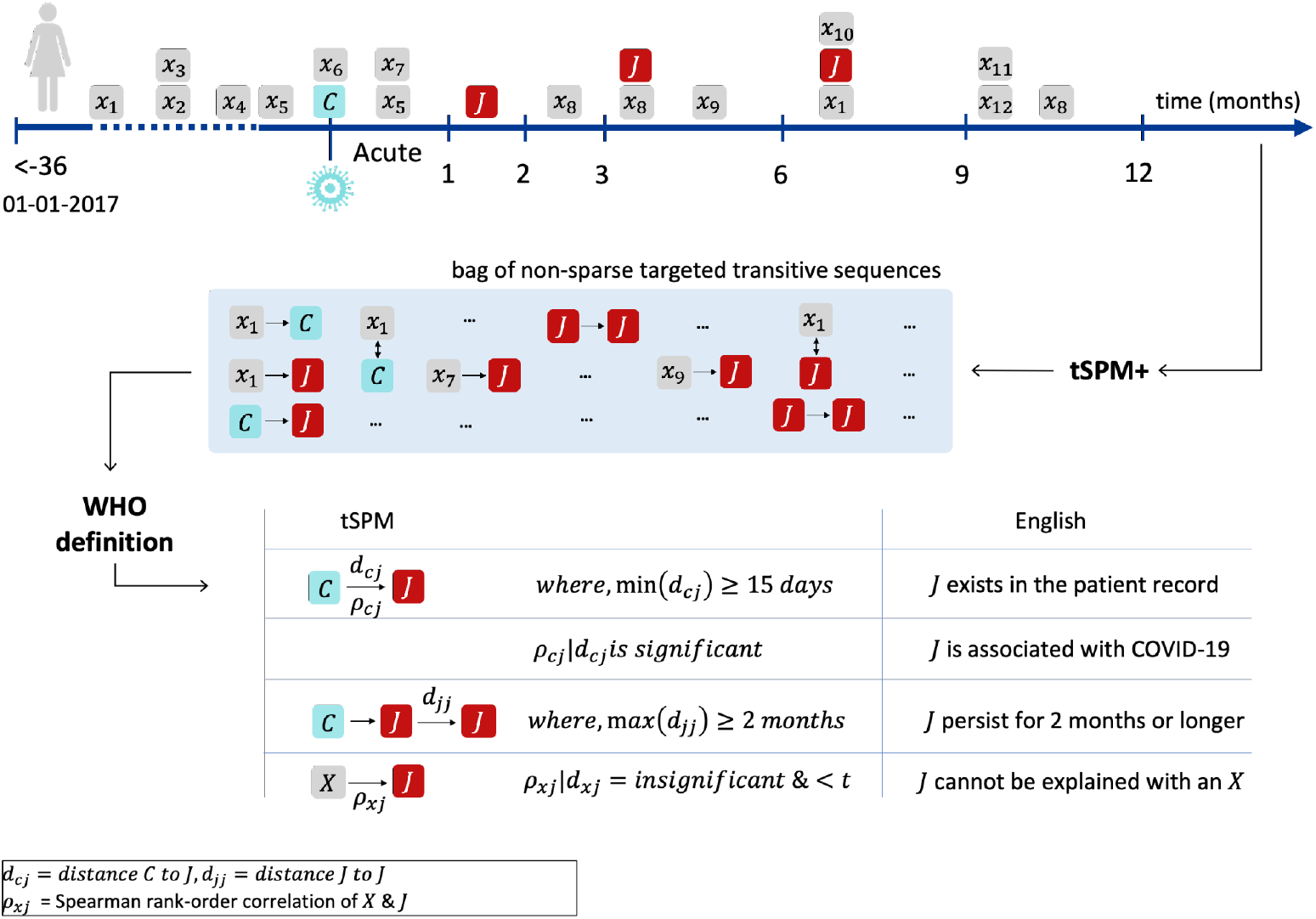
Identifying PASC with the tSPM+ algorithm and WHO definition. After temporarily ordering clinical records, we mine transitive sequences. To meet the WHO definition for PASC, a candidate problem, *J*, must have multiple sequences following a COVID-19 infection, have a significant temporal association with COVID-19, and have no temporal association with prior conditions.

### Chart Reviews

We performed an extensive chart review to explore the unstructured data in the clinical notes and to label patients who truly have PASC. The inclusion criteria for chart reviews included patients with at least an ICD-10 code U09.9 withing the study period. Eight clinical faculty developed chart review guidelines, and two clinical nurses trained in this specific chart review process reviewed the charts between April and November 2023 (see Appendix 2 for more details).

### Validation and tuning

To validate the results and estimate cut-off thresholds for the exclusion by temporal association (second step in PASC definition), we leveraged 309 chart-reviewed patients with confirmed PASC and computed positive predictive values (PPV) via bootstrapping. We first identified a cut-off threshold for each PASC that maximized PPV and then aimed to minimize false positive detection rates (type I error); we utilized clinical expertise to identify a subset of the *J* from the long-haulers list that may not contribute to the PPV. Details of the validation and tuning procedures are provided in Appendix 2.

### Statistical analysis

We computed the Spearman rank-order correlations^67^ and p-values with the Holm–Bonferroni^68^ adjustment for multiple comparisons to measure temporal associations rho (***ρ***) between clinical concepts in the four temporal buckets. We fitted the logistic regression models with a binomial logit link function to separately evaluate the development of organ specific PASC, using age, sex, race, ethnicity, and Charlson’s comorbidity score as predictors. A significance level of 0.05 was used to evaluate statistical significance.

## Result

There were 85,364 COVID-19 cases, 170,497 (matched 1:2) post-pandemic controls, and 39,817 participants in the pre-pandemic control group who met our EHR longitudinal continuity threshold. The COVID-19 group had a mean age of 53.6 years (s.d.: 17.2), and 62.6 percent of participants were female. The post- and pre-pandemic control groups had mean ages of 53.7 years (s.d.: 17.2) and 54.7 years (s.d.: 17.8), and 62.5 percent and 63.2 percent of participants were female, respectively (Table 1). All patients with an infection were followed up for 12 months after their last infection episode.

We evaluated 434 CCSR categories for PASC, 66 of which remained in the final output after applying the diagnosis of exclusion (Table 2S, appendix). For benchmarking, we reviewed clinical charts from 862 randomly selected patients with a U09.9 ICD-10 diagnosis code. In 671 (77.8 percent), the PASC diagnosis code was true. Figure 1S (appendix) illustrates the bootstrap validation estimates of the positive predictive values across the cumulative correlations. After minimizing the type I error, the PPV/precision of our PASC phenotyping algorithm was 79.9 percent – i.e., a 2.7 percent improvement in precision over U09.9. Figure 2S (appendix) illustrates an entry from the long-haulers list output, representing a hypothetical PASC patient.

**Table 2.** Summary statistics comparing long-haulers with non-long-haulers.

|  | Long-haulers<br>(N=24,360) | non-long-haulers*<br>(N=61,004) |
| --- | --- | --- |
| **Age** |  |  |
| Mean (SD) | 57.1 (17.4) | 52.2 (16.9) |
| Median [Min, Max] | 58.0 [19.0, 106] | 53.0 [19.0, 104] |
| **Age Group** |  |  |
| <45 years old | 6,389 (26.2%) | 21,792 (35.7%) |
| 45-65 years old | 9,595 (39.4%) | 24,721 (40.5%) |
| >65 years old | 8,376 (34.4%) | 14,491 (23.8%) |
| **Sex** |  |  |
| Male | 8,653 (35.5%) | 23,319 (38.2%) |
| Female | 15,707 (64.5%) | 37,685 (61.8%) |
| **Race** |  |  |
| White | 17,385 (71.4%) | 43,550 (71.4%) |
| Black | 2,538 (10.4%) | 5,870 (9.6%) |
| Asian | 700 (2.9%) | 2,233 (3.7%) |
| Other | 3,059 (12.6%) | 7,184 (11.8%) |
| Unknown | 678 (2.8%) | 2,167 (3.6%) |
| **Hispanic** | 1,612 (6.6%) | 4,498 (7.4%) |
| **Charlson Index\*\*** |  |  |
| Mean (SD) | 3.1 (2.9) | 1.8 (2.2) |
| Median [Min, Max] | 2.0 [0, 19.0] | 1.0 [0, 18.0] |
| **Charlson 10-year probability of survival** |  |  |
| Mean (SD) | 66.7 (37.0) | 81.8 (27.7) |
| Median [Min, Max] | 90.1 [0, 98.3] | 95.9 [0, 98.3] |

\* the COVID-19 patients who did not develop long terms sequelae of COVID-19
\*\* calculated before first episode of COVID-19

In the study period, 6,340 patients had a record of U09.9 in the MGB clinical data repository, though only 3,970 (62.6 percent) had a positive COVID-19 record. 80.1 percent of patients with a U09.9 diagnosis record were White, 4.83 percent were Black or African American, 2.7 percent were Hispanic, and 67.5 percent were Female. Over 62 percent (3,970) of the patients with a U09.9 record had a record indicating COVID-19 infection (diagnosis code or positive PCR test).

The precision PASC phenotyping algorithm identified 24,360 patients (28.5 percent of the cases) as long-haulers, having at least one PASC. According to our algorithm, over 13 percent of COVID-19 patients had more than one PASC, with four percent afflicted by more than three. 71.4 percent of long-haulers were White, 10.4 percent Black, 6.6 percent Hispanic, and 64.5 percent were Female (Table 2). Of the patients with a U09.9 diagnosis code, 2,104 had both medical record(s) indicating COVID-19 infection(s) and met our data continuity criteria. Our algorithm picked up 73.8 percent of these patients.

In Figure 3 and Figure 4, we plot the distribution of PASC by organ (see Table 3S in the appendix for more details). Of the 85,364 COVID-19 patients (cases), 10,044 (11.8 percent of the cases and 41.2 percent of the long-haulers) experienced systemic post-COVID sequelae, including edema, generalized pain, sleep disorders, change in weight or nutrition, and malaise and fatigue. More rare systemic complications included dizziness, dysphonia, hyperhidrosis, and sexual dysfunction in the form of low libido. About 7,000 patients had cardiovascular PASC, including palpitations, changes in heart rate (tachy/bradycardia), chest pain, dysrhythmia, changes in blood pressure (hyper/hypotension), and more rarely coronary atherosclerosis, heart failure, and myocarditis.

**Figure 3.**
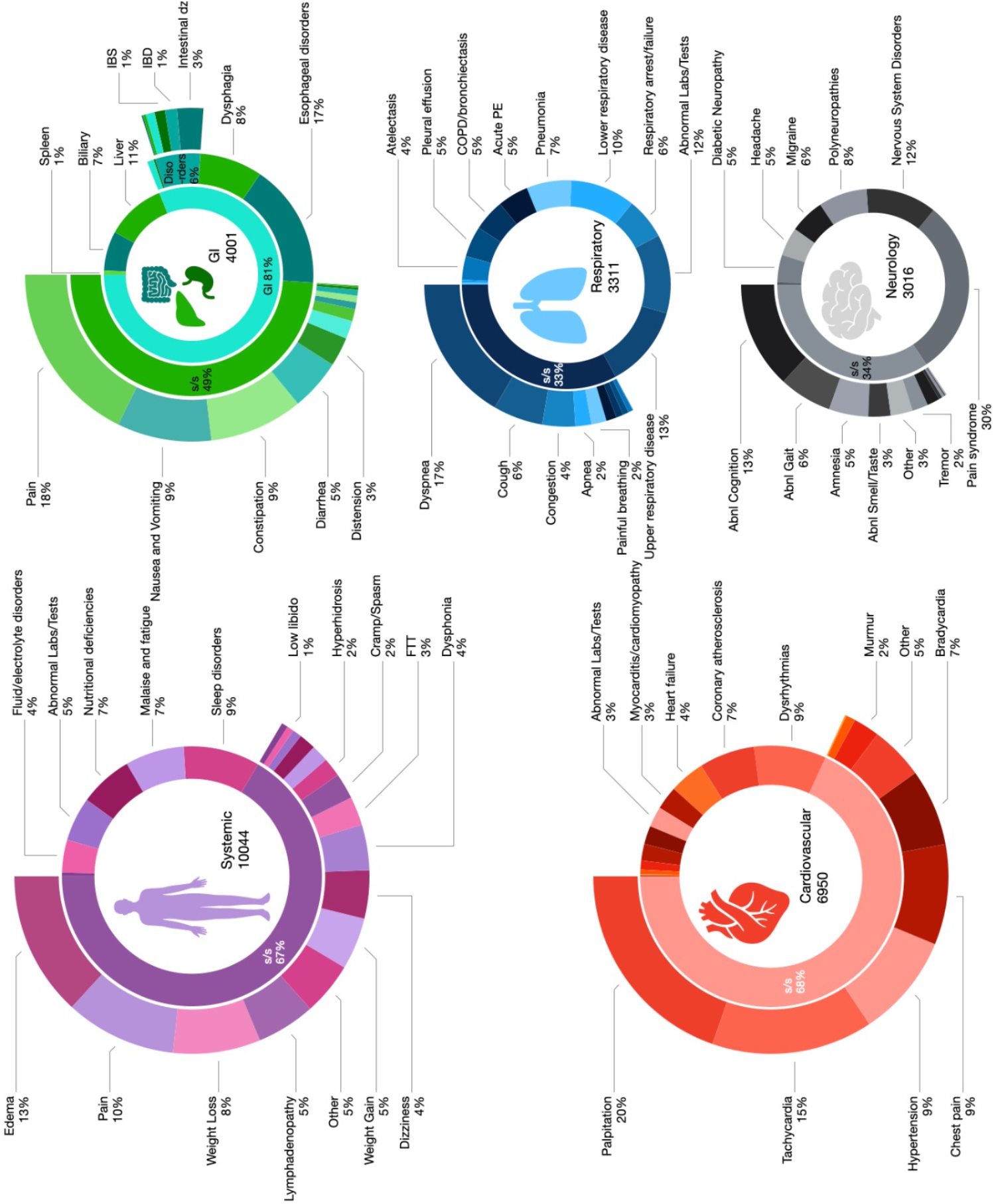
Distribution of PASC by organ. This figure focuses on the systemic, cardiovascular, neurology, respiratory, and gastrointestinal (GI) PASC. Underlying issues are presented as the percentage of patients afflicted in each category. Uses the following acronyms: s/s: signs and symptoms; Abnl: Abnormal; FTT: failure to thrive; IBS: inflammatory bowel syndrome; IBD: inflammatory bowel disease; dz: disease

**Figure 4.**
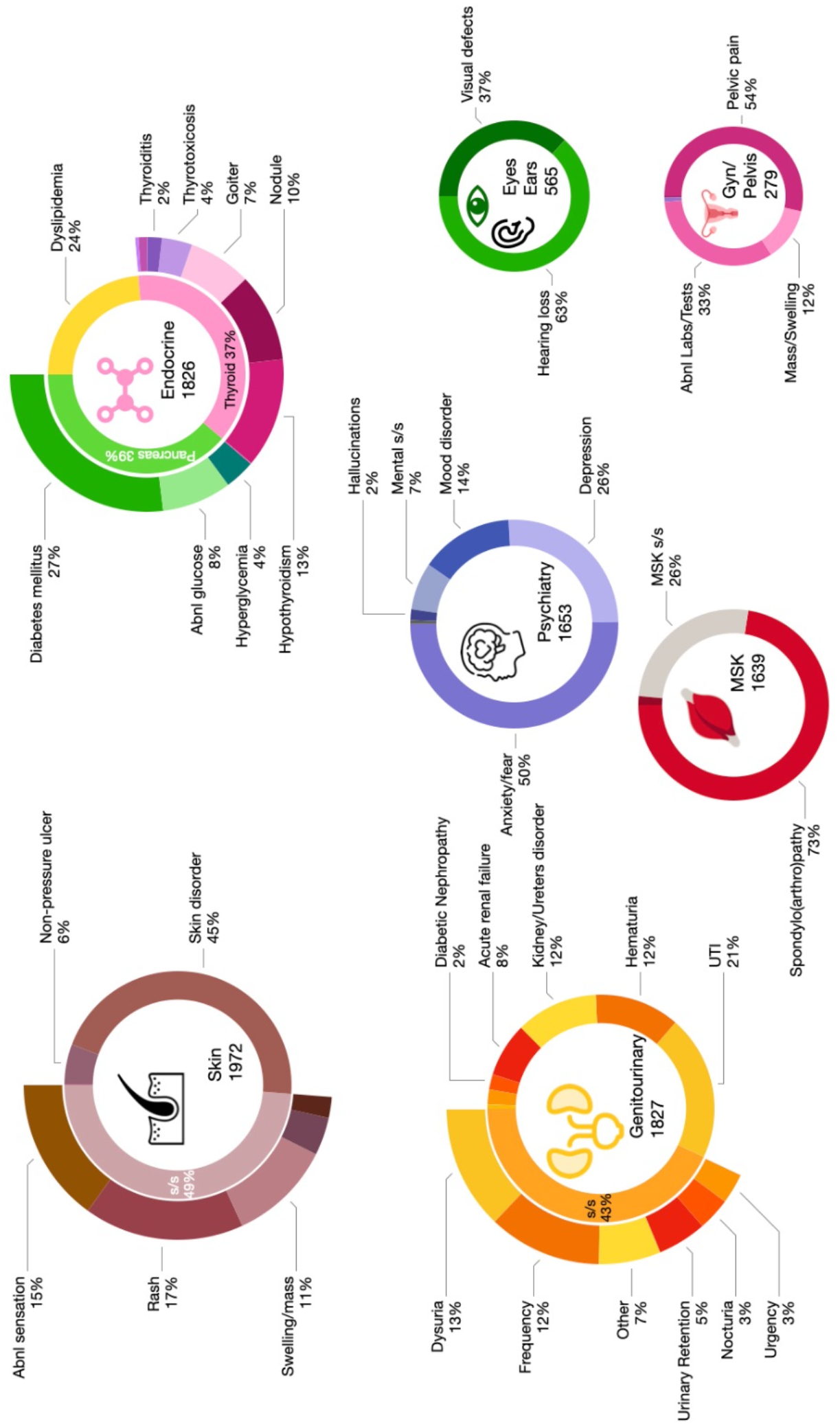
Distribution of PASC by organ. This figure focuses on the genitourinary, skin, endocrine, psychiatry, eyes and ears, musculoskeletal (MSK), and gynecology (GYN)/pelvis PASC. Underlying issues are presented as the percentage of patients afflicted in each category. Uses the following acronyms: s/s: signs and symptoms; Abnl: Abnormal.

Over 4,000 patients (4.7 percent cases and 16.4 percent of long-haulers) experienced long-term gastrointestinal problems, including prolonged abdominal pain, nausea, and vomiting, changes in bowel habits (constipation/diarrhea), esophageal disorders and dysphagia, as well as biliary and liver-associated symptoms. Less than 1 in 25 patients with a past SARS-CoV-2 infection, comprising 13.6 percent of long-haulers, experienced respiratory sequelae, including upper/lower respiratory disease, shortness of breath, cough, pneumonia, respiratory arrest, pulmonary embolism, COPD exacerbations, pleural effusion or atelectasis. About 3,000 patients experienced neurological PASC, including pain syndromes and polyneuropathies, nervous system disorders, abnormal cognition, gait, smell and taste, and headache.

Skin PASC was observed in 1,972 patients (8.1 percent of long haulers), including skin disorders, prolonged rashes, paresthesias, and swelling. Genitourinary PASC affected 1,827 patients including urinary tract infections, hematuria, renal disorders and sometimes failure, and other genitourinary signs and symptoms. Similarly, endocrine sequelae affected 1,826 patients, including problems with lipid, glucose (i.e., diabetes), and thyroid hormone (i.e., hypothyroidism) regulations. Mental health sequelae, including anxiety, fear-related disorders, and depression, affected 1,653 patients. Musculoskeletal PASC, including spondylopathy or arthropathy, was observed in 1,639 patients. More rarely but still notably, around 1 percent of long-haulers experienced visual or hearing loss and gynecological or pelvic PASC.

After correcting demographics and comorbidities (Figure 5), we found that women have significantly higher odds of developing systemic, GI, neurological, skin, mental health-related, musculoskeletal, and gynecological PASC than men. Odds of respiratory, renal/GU, endocrine, and eyes and ears PASC were not significantly different at p-value < 0.05 (ORs in Table 4S, appendix). We also found statistically significant reduced odds among Hispanic patients in 50 percent of the organ categories. Asians were more likely to develop endocrine PASC and less likely to have cardiovascular and systemic PASC than Whites. The increased odds of PASC among Black patients were statistically significant in skin, renal/GU, neurologic, cardiovascular, and systemic organs. Gynecological, skin, and psychiatric PASC had considerably higher odds among patients under 45 of age, whereas PASC problems related to eyes and ears, endocrine musculoskeletal, and renal/GU systems were more likely among older COVID-19 patients.

**Figure 5.**
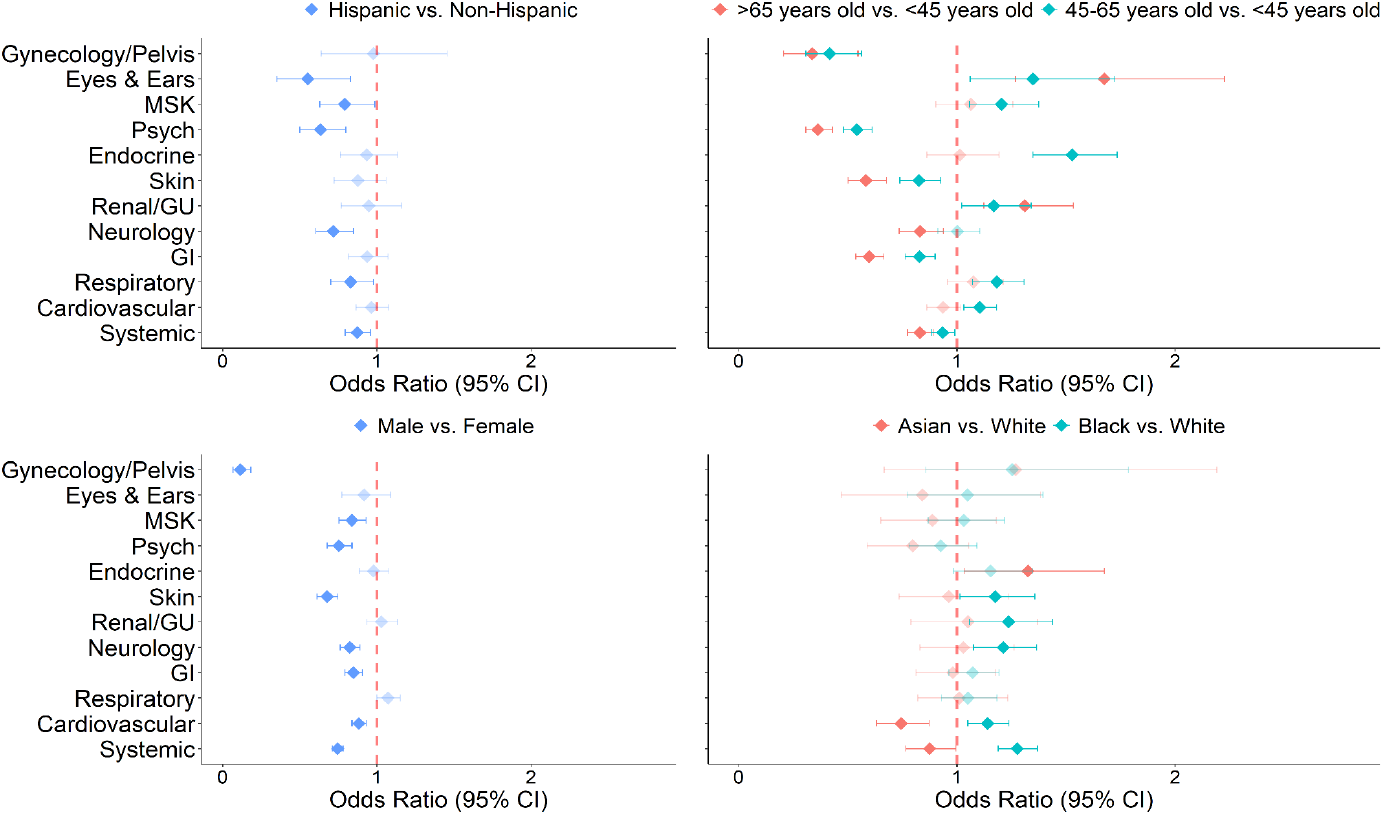
Odds of developing PASC by organ and demographics.

Most sequelae emerge within the first three months following COVID-19 infection and persist for two months or longer (Figure 6). This temporal pattern is more pronounced in respiratory, cardiovascular, neurology, general, and GI problems. Specific sequelae, such as respiratory or renal failure, are more likely to occur right after the acute phase of the infection. In contrast, weight gain, diabetic nephropathy, or hypertensive heart disease tend to occur later on. Some PASC, such as autoimmune disorders, seem to be bimodal, increasing in frequency both immediately after and four months after Covid infection. Other sequelae seem to have a steadily uniform frequency in occurrence over the first six months and then decrease afterward, such as pain syndromes or mental health signs and symptoms.

**Figure 6.**
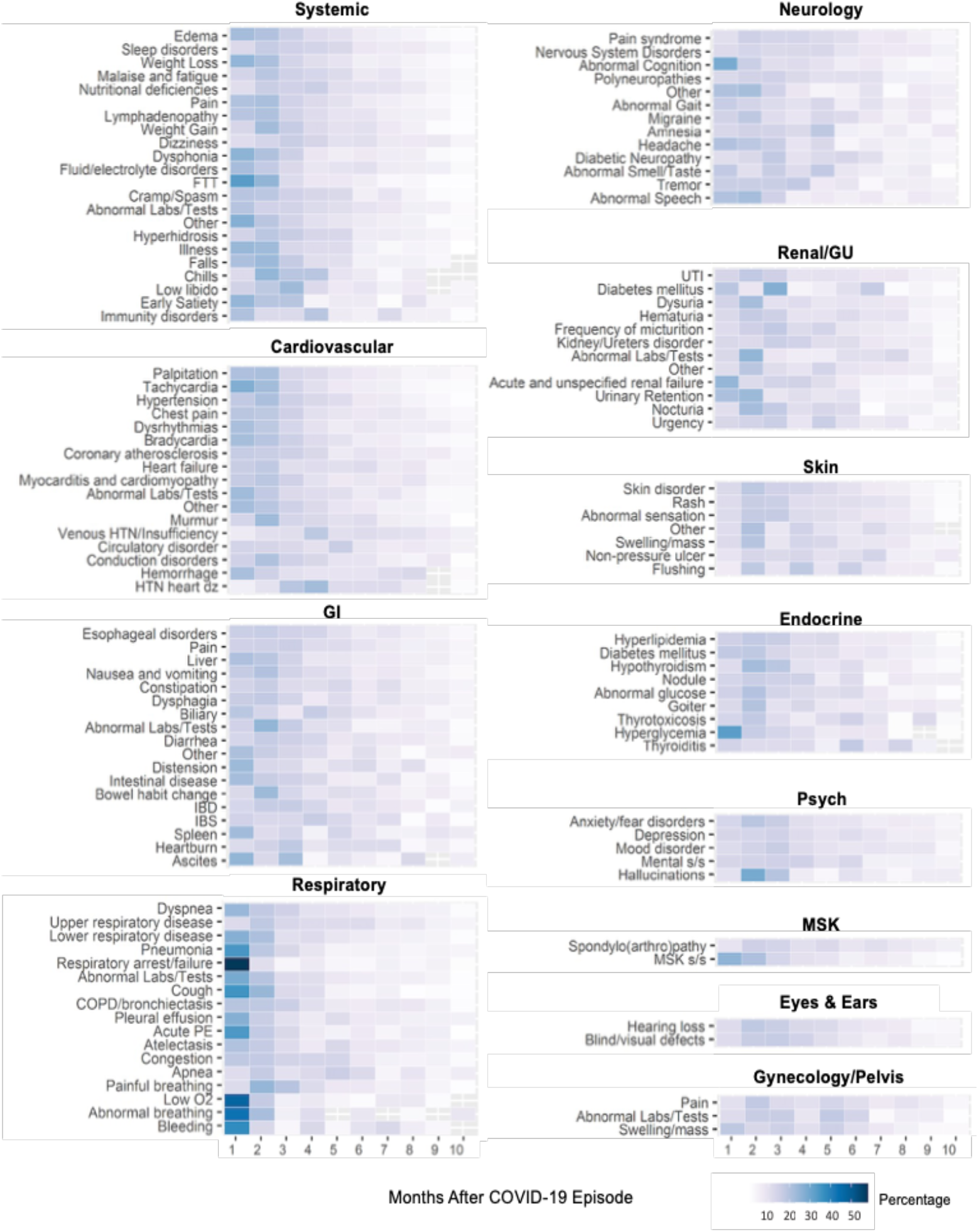
Temporal presentation of the PASC over time.

## Discussion

We presented a precision phenotyping algorithm for identifying patients with post-acute sequelae of COVID-19 (PASC). Compared to relying solely on the U09.9 diagnosis code to identify Long COVID patients, our method boasts superior precision, accurately gauges the prevalence of PASC without underestimating Long COVID, and exhibits less bias across demographic groups.

Standard risk studies identifying health outcomes with higher relative risks among COVID-19 patients are insufficient for identifying long-haulers. PASC can be explained in some patients by a prior comorbidity and/or procedure. Relying on the WHO’s definition, our approach takes the prevalent PASC risk studies to the next level by adding an extra step: diagnosis of exclusion. From all patients infected with SARS-CoV-2, our algorithm distinguishes those with PASC that were not attributable to another documented pre-existing condition.

The ICD-10 code currently in use for PASC patients, U09.9, is not precise enough in identifying affected patients. Zhang et al. (2023) showed that relying on U09.9 as a stand-in for PASC can be unreliable, with its positive predictive value (PPV)/precision fluctuating between 40 and 65 percent, depending on the PASC reference definition.^51^ Our chart reviews of nearly 900 patients indicated a PPV of 77.8 percent for the U09.9 diagnosis code. As such, our precision PASC phenotyping algorithm provided a 2.7 percent improvement in precision (79.9 percent). This demonstrates that the accuracy of our algorithm is at least equal to the only available diagnosis code that is supposed to be used in clinical care to identify PASC patients.

Our algorithm also outperformed the U09.9 in providing a more accurate estimate of Long COVID prevalence, suggesting that the latter may be falling short in capturing the true scope of the condition. Based on estimates from the National Center for Health Statistics^69^ derived from data spanning June 1, 2022, to October 2, 2023, the prevalence of PASC in Massachusetts is 24.0 percent. Our algorithm indicated a raw estimate of 28.5 percent. With a positive predictive value (PPV)/precision of 79.9 percent, our adjusted prevalence estimate is 22.8 percent (28.5 percent * 79.9 percent). Meanwhile, in the study period, 6,340 patients had a record of U09.9 in the MGB clinical data repository, which, considering the 77.8 percent precision, would lead to an adjusted estimated number of under 5,000 PASC patients.

In general, bias is a significant issue in EHR diagnosis codes. It has been reported that the demographic composition of patients diagnosed with U09.9 leans toward females, White, and non-Hispanic patients.^48^ We found similar biases; 80.1 percent of patients with a U09.9 diagnosis code were White, 4.83 percent were Black or African American, 2.7 percent Hispanic, and 67.5 percent were Female. According to the US Census, 69.6 percent of the Massachusetts population is White, 51 percent is Female, 9.5 percent is Black or African American, and 13.1 percent is Hispanic.^80^ Our algorithm also provides a more unbiased distribution of PASC patients across race, gender, and ethnicity compared to the U09.9 diagnosis code. 71.4 percent of long-haulers identified by our precision phenotyping algorithm were White, 10.4 percent were Black or African American, 6.6 percent were Hispanic, and 64.5 percent were Female.

We also provided an in-depth analysis outlining the clinical attributes, encompassing identified lingering effects by organ, comorbidity profiles, and temporal differences in the risk of PASC. Of the 24,360 long-haulers in our cases, less than half had multiple PASC, which could be in the same infection episode or subsequent infections. The approach to identifying cohorts with PASC offers the highest precision to date. For example, we identified PASC linked with different episodes of COVID-19 infections. We found that having a prior PASC increases the chances of having more PASC in subsequent infections. This could be due to long-haulers’ inability to mount an appropriate, timely immune response to clear the COVID-19 infection each time, leading to greater susceptibility to developing PASC in subsequent infections.

Most of the post-acute sequelae of COVID-19 we identified in this study (systemic symptoms including malaise and fatigue, sleep problems) have also been reported by prior research. However, our algorithm also allowed us to go a step further by identifying more rare PASC such as vision or hearing loss, loss of libido resulting in sexual dysfunction, gynecological complications, diabetic complications in various organs such as diabetic nephropathy, neuropathy, or vasculopathy. Further, our estimates are more realistic as we exclude sequelae that can be explained at the patient level. For example, we find that only 1 percent of our COVID-19 cases suffered from long-term malaise and fatigue that can be attributed to a SARS-CoV-2 infection episode, compared to much higher rates reported in other studies.^70,71^

Our precision phenotyping enabled us to discover statistically significant differences in the odds of developing PASC among racial and ethnic groups and across organ systems, which are not well studied. For example, gynecological, skin, and psychiatric PASC had considerably higher odds among patients under 45, whereas PASC problems related to eyes and ears, endocrine musculoskeletal, and renal/GU systems were more likely among older COVID-19 patients. Women had higher odds of PASC in 8 organs than men. Asian (vs. White) and Hispanic (vs. non-Hispanic) patients had lower odds of developing PASC (mainly in the cardiovascular system), and Black (vs. White) patients had greater odds of PASC, regardless of comorbidity and age. Black patients’ higher odds were statistically primarily in the skin, renal/GU, neurologic, cardiovascular, and systemic organs.

Using structured clinical data from real-world settings for studying PASC signs and symptoms may be limiting, as this information is often better documented in clinical notes. As we demonstrated in this study, structured diagnosis codes capture an array of signs and symptoms. We picked the CCSR categories to work with a manageable and clinically meaningful grouping of conditions, signs, and symptoms. This allowed us to reduce the computational costs of running our algorithms and facilitate implementation in diverse settings. We traced the CCSR categories to the data entries for further clinical interpretations and analyses.

Our reliance on structured data may have resulted in an underestimation of PASC. However, it has been shown that the transitive sequential patterns of the events stored in clinical data can elevate signal detection from structured data, compensating for the possible loss of information.^79^ Future studies can incorporate timestamped signs and symptoms from clinical notes.

Another limitation of this study is that we did not capture the possible worsening of a prior condition, which could be characterized as PASC. For example, COVID-19 could lead to prolonged COPD exacerbation; however, if the patient had prior episodes of COPD exacerbations prior to the Covid infection, this likely has been removed per our diagnosis of exclusion. Identification of such possible PASC will be complex and require the inclusion of information on the severity of records over time. Finally, we separated COVID-19 variants using the date of infection rather than genetic data, thus, interpretations of the variant analysis should be cautiously approached.

Implementing the algorithm to curate similar cohorts depends on the availability of a true or estimated date of infection. This is a limitation, given that many do not test for COVID-19 anymore. To overcome this limitation, future research can build up on this work to develop precision definitions for PASC phenotypes, which can then be utilized retrospectively to develop “postdiction”^57^ algorithms for identifying who may have had SARS-CoV-2 infections in the past.

The attention mechanism we developed in this study relied on the bootstrap tuning approach for identifying thresholds for exclusion by temporal association. Our tuning approach only relied on optimizing positive predictive values as we could only validate cases with PASC. Future work can further expand this concept by evaluating the generalizability of exclusion thresholds for different concepts and temporal windows, increasing the validation samples, and incorporating additional validation criteria, such as negative predictive value.

With the programs and data we offer alongside this publication, this cohort can now be curated in any healthcare system with reliable longitudinal diagnosis and procedure data on their patients. Access to large cohorts of long-haulers enriched with the precision offered by this algorithm will offer unprecedented opportunities to stratify patients who are at risk for post-COVID sequelae, identify genetic risk factors for PASC, study the possible impacts of therapeutics and immunizations, and diversify recruitment for clinical studies on post-acute sequelae of COVID-19.

Compared to the conventional U09.9 diagnosis code, our method for identifying PASC boasts superior precision and exhibits less bias, accurately gauging the prevalence of this condition without downplaying its significance, offering a more nuanced understanding of Long COVID patients. The comprehensive PASC cohort resulting from our precision phenotyping algorithm will enable deep dives into the multifaceted expressions of Long COVID through genetic, metabolomic, and clinical inquiries bolstered by robust statistical prowess, which surpasses the constraints of earlier PASC cohort studies due to limited size and outcome data.

## Data Availability

Clinical data used in the study are not available publicly. All non-clinical data produced are available online at https://github.com/clai-group/long_covid_ai_scripts/pkgs/container/post_covid_ai_scripts

## Data Sharing

The computer codes can be accessed at https://github.com/clai-group/long_covid_ai_scripts or via docker at https://github.com/clai-group/long_covid_ai_scripts/pkgs/container/post_covid_ai_scripts

## Ethics approval

Use of patient data in this study was approved by the Mass General Brigham Institutional Review Board (protocol 2020P001063).

## Acknowledgments

This study has been supported by grants from the National Institutes of Health, National Institute of Allergy and Infectious Diseases (NIAID) R01AI165535, National Heart, Lung, and Blood Institute (NHLBI) OT2HL161847, and National Center for Advancing Translational Sciences (NCATS) UL1 TR003167, UL1 TR001881, and U24TR004111. J.Hügel’s work was partially funded by a fellowship within the IFI programme of the German Academic Exchange Service (DAAD) and by the Federal Ministry of Education and Research (BMBF) as well by the German Research Foundation (426671079).

